# Prevalence and National Patterns of Commonly Prescribed Antidepressants and other Psychotropic Medications to patients with treatment-resistant depression in France

**DOI:** 10.1101/2024.01.18.24301467

**Authors:** Vimont Alexandre, Biscond Margot, Leleu Henri, Marina Sanchez-Rico², Hoertel Nicolas, Llorca Pierre-Michel

**Author notes:** **Funding:** This study was fully funded by Public Health Expertise. **Author Contributions:** Conceptualization: A.V, M.B, N.H and P.M.L. Statistical analyses: A.V, M.B. Writing—Original Draft Preparation: A.V, M.B; Writing—Review and Editing: A.V, M.B, N.H and P.M.L. All authors have read and agreed to the published version of the manuscript. **Informed Consent Statement: Not applicable**. **Data Availability Statement** Data are not publicly available. **Conflict of interest:** Pierre-Michel Llorca has received personal fees and/or non-financial support from Eisai, Ethypharm, Janssen, Lundbeck, MSD, Neuraxpharm, Novartis, Otsuka, Roche, Rovi, and Sanofi. The other authors declare no conflicts of interest.

## Abstract

**Background:** Prevalence of Treatment-Resistant Depression (TRD) varied widely across studies due to heterogeneous definitions. Several treatment strategies exist to manage patients with TRD but evidence from real-world data is scarce. Investigating their use in real-life settings is important to understand national prescribing practices and to refine prevalence estimation.

**Method:** All adult patients (≥ 18 years) with a TRD episode for the year 2019 were identified in a sample of four French regions accounting for 27% of national individuals. After exclusion of patients with psychotic or bipolar disorders, Parkinson’s disease, and dementia, TRD was defined by i/ 3 successive sequences of different antidepressants (AD), or ii/ the dispensing of several different AD together, or iii/ an AD with a potentiator (lithium, antiepileptic drugs, or antipsychotic drugs) over the same treatment period. The prevalence rate was estimated for the year 2019 and treatment patterns were described by treatment class and molecule.

**Results:** For the year 2019, 66,810 patients were identified with TRD, accounting for 23.9% of all patients treated for depression. The mean age was 56 years (±15.9) with 63.7% of women. Standardized prevalence of TRD was estimated at 35.1 per 10 000 patients, and 25.8 per 10,000 patients when excluding patients probably treated for another primary diagnosis than depression. Association of an AD with an antipsychotic was the most frequently used strategy, with SSRIs and second-generation antipsychotics being the most often prescribed.

**Conclusion:** This study provides robust population-based estimates of the prevalence of TRD in the French population. Description of treatment patterns highlight the widespread use of second-generation antipsychotics as potentiator of antidepressants.

## Introduction

Major depressive disorder (MDD) is highly prevalent in the global population, affecting 5.1% of females and 3.6% of men, varying according to age and region (1,2). In France, 1.7% to 7.5% of the population present with MDD (3) and 12.5% of adults report having experienced a major depressive episode (MDE) within the last 12 months (4). Depression is a major public health priority since it is associated with a high economic burden (5), social consequences (6) and increased risk of suicide (7,8), especially in adolescents and young adults and in older adults (9–11).

Treatment options for MDE include psychotherapy, pharmacotherapy, and neuromodulation strategies such as electroconvulsive therapy (ECT) and repeated transcranial magnetic stimulation (rTMS) (12). Although treatment recommendations support decision making to physicians to improve the chances of achieving a response (13), a substantial proportion of patients still do not achieve remission from depression (14). For example, antidepressant monotherapy produces an approximately 60% response rate and only 40% remission rate (15).

A consensual definition states that patients who failed to respond adequately after the consecutive use of at least two antidepressants (ADs) are considered treatment-resistant to depression (TRD) (13). AD could whether or not be from different pharmacological classes or being combined, but at doses and durations that would normally be effective, administered for 6 to 8 weeks to a patient who is adherent (16,17).

Recommendations suggest that sequence of pharmacotherapies may include the combination of two individual ADs in case of non-response to an antidepressant monotherapy or a combination of an AD with another medication with mood stabilizing properties such as lithium, certain antiepileptics (e.g. valproate), certain antipsychotics (e.g. quetiapine) or thyroid hormones (18). However, the use of these combinations from real-world evidence remains scarce (19,20).

Although clinical characteristics of TRD patients have been widely described (21), the epidemiologic situation and the treatment strategies used in real-world remain poorly documented since definitions of TRD were heterogeneous across studies, especially regarding the number or the type or the dose of antidepressants (AD) and the evaluation of patients’ compliance to the treatment. However, it is well known that the clinical and economic burden is amplified among TRD patients, who are associated to increased impairment and morbidity, and less likely to respond to a subsequent sequence of therapy (22).

In this study, we conducted a retrospective large-scale population-based study with the French nationwide claims database to estimate the prevalence of TRD and to examine treatment strategies used by practitioners in real-world settings.

## Methods

### Data source

Data were drawn from a sample of the French nationwide claims database (SNDS database) under the regulatory approval MLD/TDC/AR 211497 (CNIL reference number). Since the entire population from the SNDS is not made available for research (23), the sample of four French regions was used and we included adults living in these regions with at least one delivery of AD during the period 2015 to 2019. Regions included Bretagne, Normandie, Loire-Atlantique, and Grand-Est, accounting for 27% of the national population. Regions were selected based on their heterogeneity in terms of incidence rates of MDD and care-settings utilization (hospital and outpatient psychiatric wards) to increase its representativeness (24,25).

This real-world database is managed by National Health Insurance (NHI) which ensures completeness of the sample. In France, a compulsory public insurance scheme is applied to all individuals to cover the majority of costs. Information on private insurance schemes that patients subscribe to cover the complementary part was not available. The database included information on demographics, medical history, diagnoses and procedures related to in-hospital admissions, prescriptions, laboratory assays, and date of death (23).

The database offers different alternatives to identify chronic diseases. First, long-term disease (LTD) coverage for all medical expenses is always related to some chronic diseases (LTD), such as MDD or dementia. Second, hospital diagnoses include primary and secondary diagnoses coded according to the 10^th^ revision of the International Classification of Diseases (ICD-10). Third, pharmacy claims provide evidence for treatment sequencing and adherence in the scope of their approved indications.

### Study design

This study included all adults with an episode of TRD identified between 1 January 2019 and 31 December 2019. Identification of TRD was based on outpatient deliveries of AD and its potentiators, complemented by other psychotic diagnoses identified either through LTD coverage either during inpatient admissions. Patients with ongoing LTD or history of admission for chronic psychotic disorder, bipolar affective disorder, Parkinson’s disease or dementia within the 5-year study period were excluded (Codes in **Supplementary file 1**). These chronic diseases were essentially identified through LTD and admission diagnoses. Anxiety disorders used in sensibility analyses were captured with co-treatment associations, essentially with anxiolytics, since the database did not allow distinction between isolated and characterized episodes.

TRD episodes during the year 2019 were identified using the standard definition of TRD and the recommended treatment strategies in France (13). TRD was defined by the failure of 2 AD-based treatment strategy (**Supplementary file 2**), including AD monotherapy, combination of 2 ADs or more, or combination of AD with a potentiator, including lithium, antiepileptic drugs without a prior history of epilepsy, antipsychotic drugs and thyroid hormones without a prior history of hypothyroidism.

An adequate treatment strategy was considered met if treatment deliveries covered at least 4-week treatment duration (duration of one prescription) followed with an adequate treatment adherence assessed by a Medication Possession Ratio (MPR) ≥ 80%. The MPR measures the proportion of days covered by treatment deliveries during the period between the first drug reimbursements of two successive treatment sequences. Because these medications are supposed to be prescribed for at minimum 9 months after remission, and often longer if it is the second episode, a recovery from TRD episode was assumed when no delivery of any AD medication was observed during a 6-month period. Thus, a subsequent TRD episode was considered when discontinuation of therapy for at least 6 months was previously observed (**Supplementary file 2**). Index date for TRD was defined at initiation of the first treatment strategy.

### Statistical analyses

A descriptive analysis of prevalent TRD patients was conducted for the year 2019. Historical period (2015–2018) was used to identified other chronic psychotic disorders and to determine the sequences of treatments. Characteristics included sex, age, duration of long-term coverage if any, duration of TRD episode (treatment duration for the current TRD episode), other psychiatric conditions, and proportion of episode initiated during the year 2019.

The prevalence was estimated for the year 2019 and extrapolated to the national level, with age and gender standardization. Prevalence was calculated by weighting the gender- and age-specific number of patients observed in the study population sample by the gender and age structure of the national population (26). A sensitivity analysis on the prevalence calculation was conducted by excluding patients being prescribed AD probably for other primary diagnosis than MDD, such as anxiety disorders, alcohol use disorder with comorbid depression, or chronic pain. These specific conditions were identified based on concomitant treatment deliveries along with AD: anxiety disorders, alcohol use disorder drugs (such as nalmefene or naltrexone), and chronic pain with opioids.

Patients assumed to be treated for a primary diagnosis of anxiety disorders were defined as patients with AD monotherapy (not potentiated) that holds an indication for anxiety disorders. Among these patients, the proportion of patients with prescribed concomitant anxiolytic or sedative treatments (benzodiazepines, H1-receptor antagonist such as hydroxyzine, and phenothiazine such as cyamemazine) were reported. Cyamemazine is a first-generation antipsychotic that holds additional approval in France for acute or long-term treatment of anxiety or aggressiveness.

Treatment strategies and molecules delivered on the last observed prescription during TRD episode for the year 2019 were reported for the overall population and stratified by group of age below and over 65 years.

## Results

### Prevalence of TRD

Among the four included regions, a total of 452,800 patients had at least one delivery of AD during the year 2019 (**Additional file 3**). Of these, 132,435 (29.3%) were excluded because of the presence of a diagnosis of chronic psychotic disorder (n=44,289, 11.1%), Parkinson’s disease (n=10,429, 2.6%), dementia (n=37,185, 9.3%), or bipolar disorder (n=39,147, 9.8%). The remaining patients were assumed to be treated for MDD (n=279,479), of which 23.9% (n=66,810) were identified with an ongoing TRD episode during 2019.

The corresponding standardized prevalence at national level was estimated at 35.4 per 10,000 patients, corresponding to 238,161 patients (31.5 per 10,000 for patients between 18 and 65, and 68.3 per 10,000 for patients over 65 years) when extrapolated to the French population.

Since it was not possible to identify precisely the exact indication of the treatment, sensitivity analyses were performed and presented in the following section by excluding patients assumed to be treated for anxiety disorders, alcohol use disorder, and chronic pain for the prevalence estimation.

### Description of the population

Patients were predominantly women (63.7%), and 29.9% (n=19,975) were more than 65 years of age (**This population-based** study supports that TRD is prevalent in the French population and highlights the frequent use of second-generation antipsychotics as potentiator of AD medication (38,3%), followed by AD monotherapy (31,9%), in real-world settings. RCTs comparing these frequent therapeutic strategies are urgently needed to reduce the suffering and the major consequences caused by this disorder.

Table 1). Most patients (64.5%) had long-term disease coverage (LTD) for depressive disorder for a median duration of 6 years (IQR: 2-13). A minority of patient had history of substance use disorder (14.1%) within the last two years (2017–2018), including alcohol use disorder (11.2%), nicotine dependence (3.4%) or other substance use disorder (4.1%). Additionally, 2.5% of patients (n=1,655) were prescribed concomitantly a treatment for alcohol use disorder in 2019 (such as nalmefene or naltrexone), anxiolytic/sedative medications (41.4%) and opioids (8.6%).

**Table 1:**
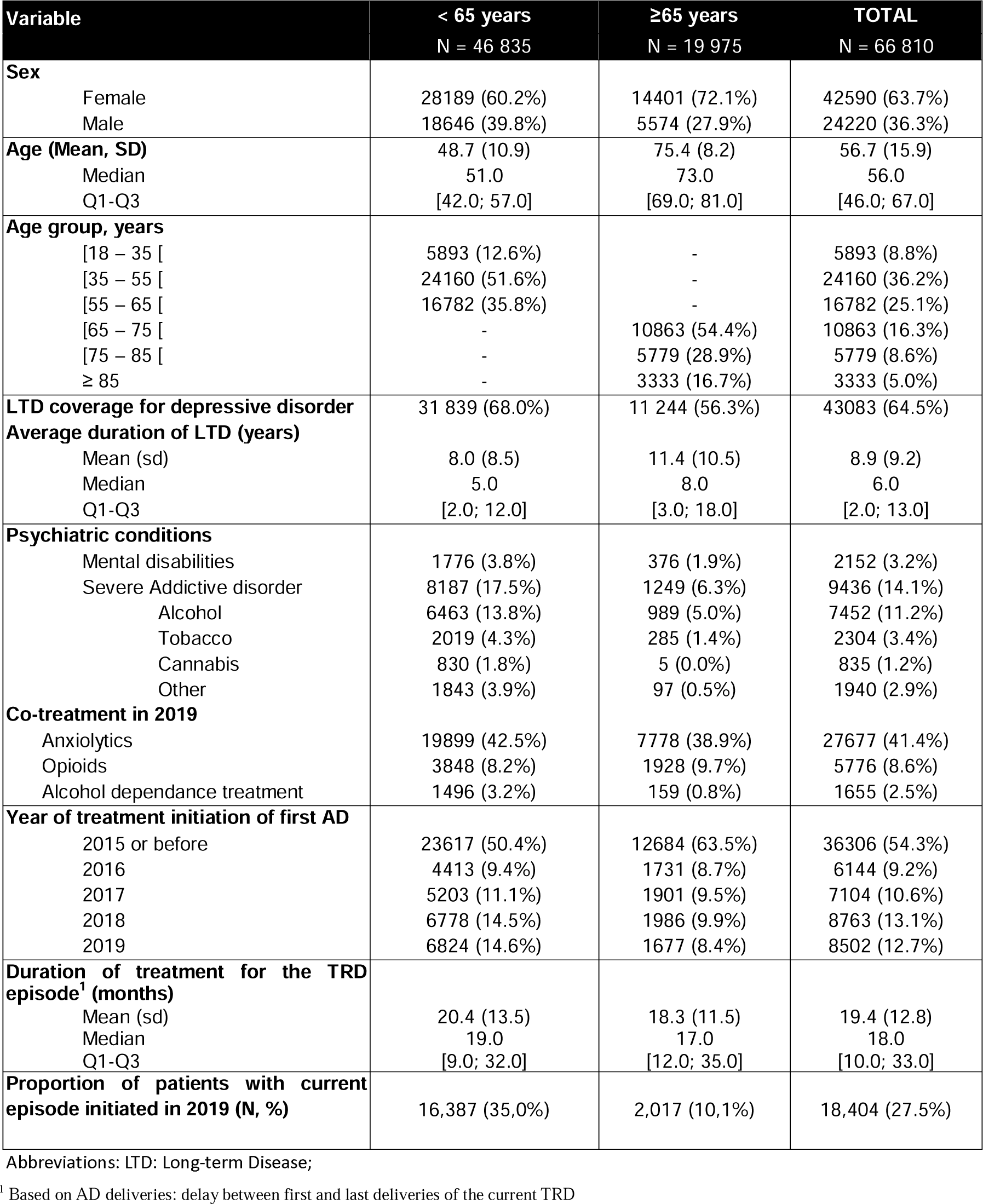
Description of TRD patients <65 years old versus. ≥ **65 years old**

| Variable | < 65 years | ≥65 years | TOTAL |
| --- | --- | --- | --- |
|  | N = 46 835 | N = 19 975 | N = 66 810 |
| **Sex** |  |  |  |
| Female | 28189 (60.2%) | 14401 (72.1%) | 42590 (63.7%) |
| Male | 18646 (39.8%) | 5574 (27.9%) | 24220 (36.3%) |
| **Age (Mean, SD)** | 48.7 (10.9) | 75.4 (8.2) | 56.7 (15.9) |
| Median | 51.0 | 73.0 | 56.0 |
| Q1-Q3 | [42.0; 57.0] | [69.0; 81.0] | [46.0; 67.0] |
| **Age group, years** |  |  |  |
| [18 – 35 [ | 5893 (12.6%) | - | 5893 (8.8%) |
| [35 – 55 [ | 24160 (51.6%) | - | 24160 (36.2%) |
| [55 – 65 [ | 16782 (35.8%) | - | 16782 (25.1%) |
| [65 – 75 [ | - | 10863 (54.4%) | 10863 (16.3%) |
| [75 – 85 [ | - | 5779 (28.9%) | 5779 (8.6%) |
| ≥ 85 | - | 3333 (16.7%) | 3333 (5.0%) |
| **LTD coverage for depressive disorder** | 31 839 (68.0%) | 11 244 (56.3%) | 43083 (64.5%) |
| **Average duration of LTD (years)** |  |  |  |
| Mean (sd) | 8.0 (8.5) | 11.4 (10.5) | 8.9 (9.2) |
| Median | 5.0 | 8.0 | 6.0 |
| Q1-Q3 | [2.0; 12.0] | [3.0; 18.0] | [2.0; 13.0] |
| **Psychiatric conditions** |  |  |  |
| Mental disabilities | 1776 (3.8%) | 376 (1.9%) | 2152 (3.2%) |
| Severe Addictive disorder | 8187 (17.5%) | 1249 (6.3%) | 9436 (14.1%) |
| Alcohol | 6463 (13.8%) | 989 (5.0%) | 7452 (11.2%) |
| Tobacco | 2019 (4.3%) | 285 (1.4%) | 2304 (3.4%) |
| Cannabis | 830 (1.8%) | 5 (0.0%) | 835 (1.2%) |
| Other | 1843 (3.9%) | 97 (0.5%) | 1940 (2.9%) |
| **Co-treatment in 2019** |  |  |  |
| Anxiolytics | 19899 (42.5%) | 7778 (38.9%) | 27677 (41.4%) |
| Opioids | 3848 (8.2%) | 1928 (9.7%) | 5776 (8.6%) |
| Alcohol dependance treatment | 1496 (3.2%) | 159 (0.8%) | 1655 (2.5%) |
| **Year of treatment initiation of first AD** |  |  |  |
| 2015 or before | 23617 (50.4%) | 12684 (63.5%) | 36306 (54.3%) |
| 2016 | 4413 (9.4%) | 1731 (8.7%) | 6144 (9.2%) |
| 2017 | 5203 (11.1%) | 1901 (9.5%) | 7104 (10.6%) |
| 2018 | 6778 (14.5%) | 1986 (9.9%) | 8763 (13.1%) |
| 2019 | 6824 (14.6%) | 1677 (8.4%) | 8502 (12.7%) |
| **Duration of treatment for the TRD episode<sup>1</sup> (months)** |  |  |  |
| Mean (sd) | 20.4 (13.5) | 18.3 (11.5) | 19.4 (12.8) |
| Median | 19.0 | 17.0 | 18.0 |
| Q1-Q3 | [9.0; 32.0] | [12.0; 35.0] | [10.0; 33.0] |
| **Proportion of patients with current episode initiated in 2019 (N, %)** | 16,387 (35,0%) | 2,017 (10,1%) | 18,404 (27.5%) |

Abbreviations: LTD: Long-term Disease;
<sup>1</sup> Based on AD deliveries: delay between first and last deliveries of the current TRD

The majority of patients were already under AD in 2015 (54.3%) and 12.7% initiated an AD in 2019 (no AD delivery within the 4-year period of 2015-2018). After excluding patients who initiated AD in 2015 or before, the median duration of treatment for the TRD episode identified in 2019 was 18 months (IQR: 10-33 months).

### Description of treatment strategies

Treatment strategies used in TRD patients in 2019 are presented in **Table 2** and **Figure 1**. Overall, 31.9% of TRD patients were treated with AD monotherapy (after failure of two different classes of AD), mainly SSRIs (11.6%). Combination therapy was used in 68.6% of TRD patients, with 58.7% receiving either 2 ADs (4.4%) or AD combined with a potentiator with mood stabilizing properties, either an antipsychotic (35.4%) or an antiepileptic (18.4%). Some patients received concomitantly 3 classes or more of ADs and a potentiator (9.4%). Overall, combination of one or several ADs associated to an antipsychotic was the most prevalent strategy (38.3% of TRD patients), followed by AD monotherapy (31.9%) after failure of two previous classes of AD.

**Figure 1:**
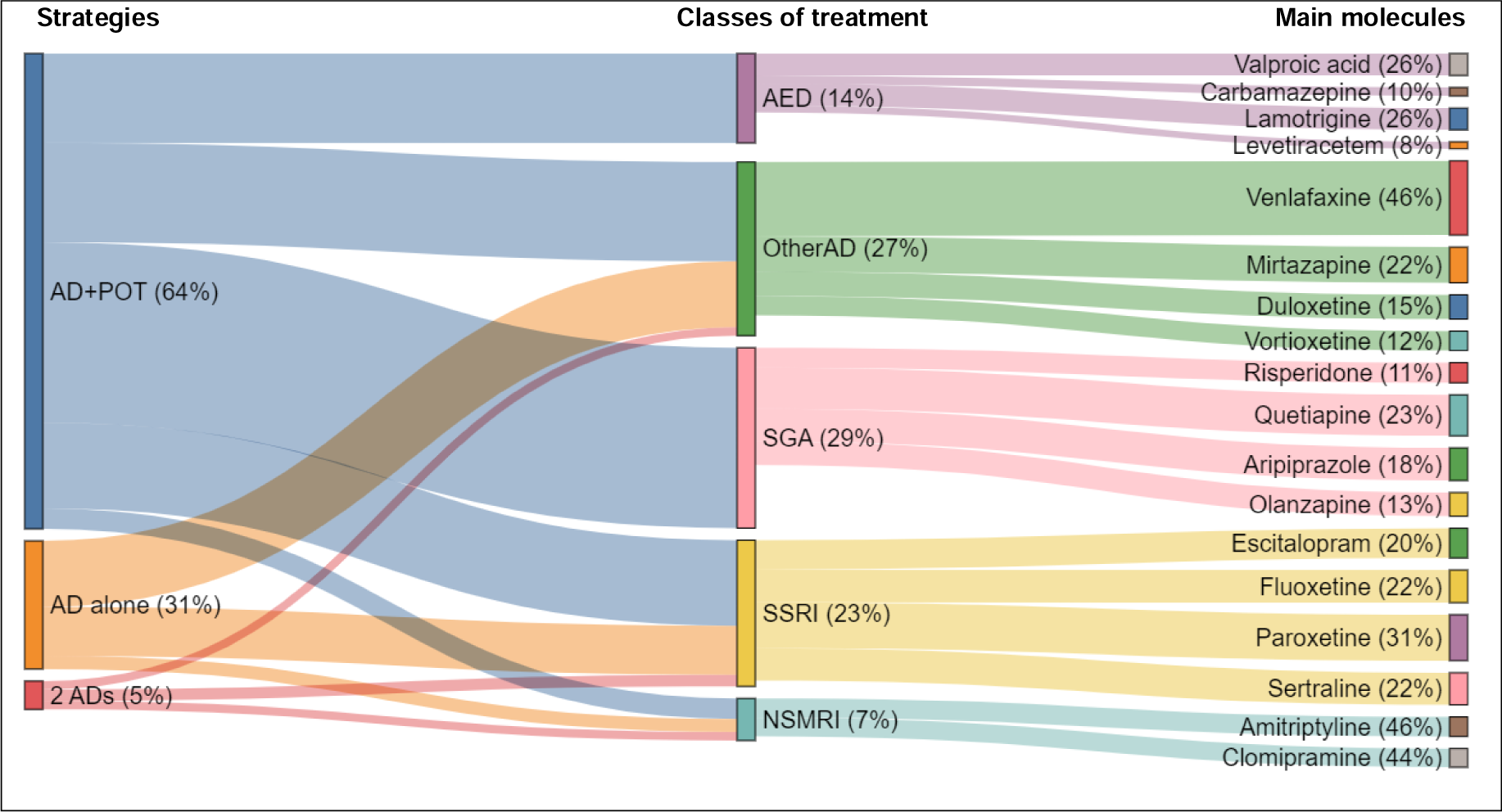
Repartition of treatment strategies, drug classes and molecules in TRD patients <65 years.

**Table 2:**
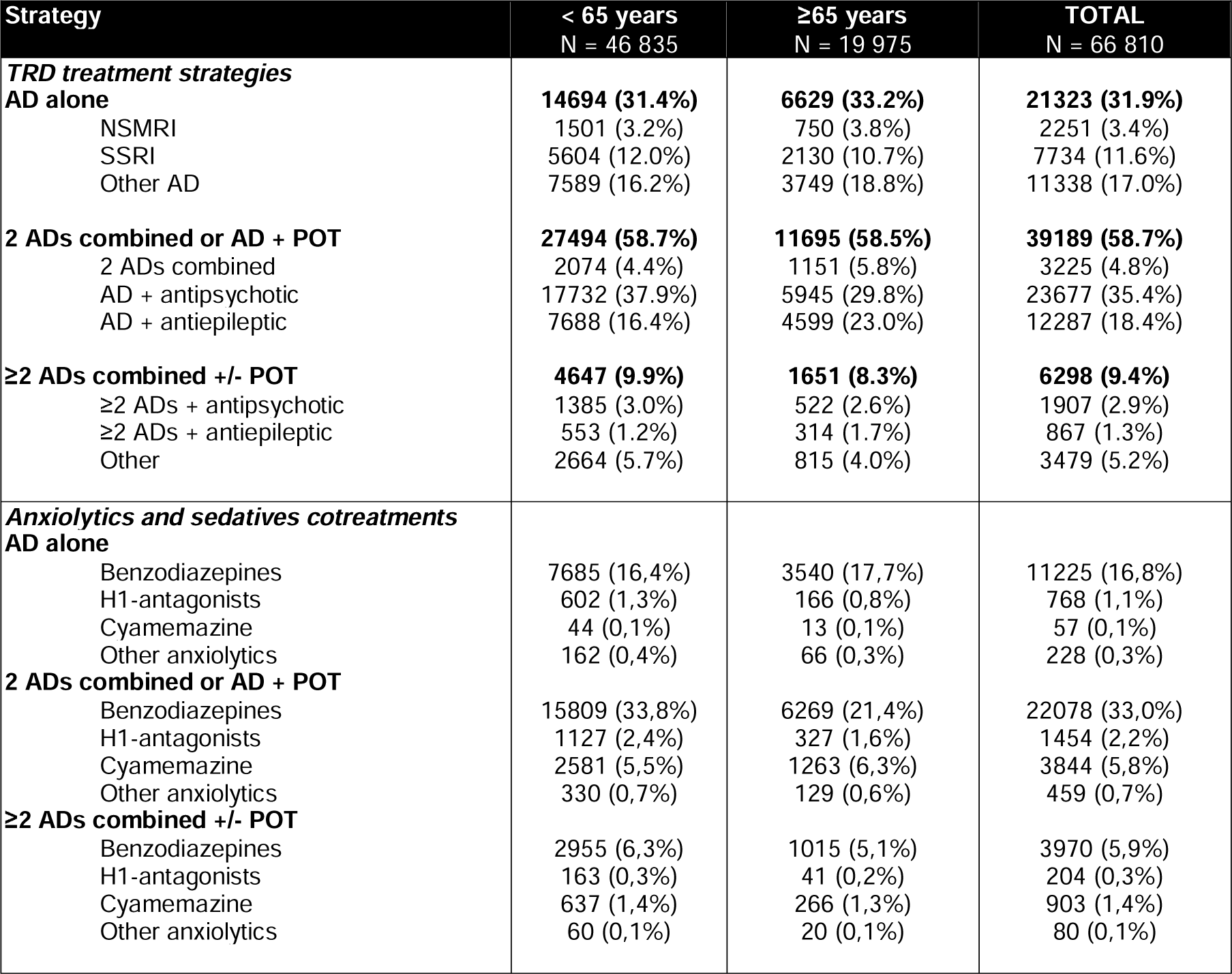
Treatment strategies and cotreatments observed at last observed delivery of TRD episode (end of follow-up)

According to patient age, treatment strategies were relatively similar except for the association of AD with an epileptic treatment which was more often prescribed in patients over 65 years old (23.0% vs 16.4%, Chi-square=253.6; p<0.0001).

Anxiolytics and sedatives co-prescribed with TRD treatment strategies are reported in **Table 2**. Overall, benzodiazepines were co-prescribed in 55.7% of patients, cyamemazine in 7.5%, and H1-receptor antagonists in 3.6%. According to patient age, proportions were similar, except for benzodiazepines co-prescribed with two Ads or AD potentiated which was less prescribed in patients over 65 years than among younger ones (21.4% vs 33.8%, Chi-square=43.33, p < 0.0001).

### Description of molecules

The proportion of each molecule used in TRD patients is presented in **Table 3**. The most prescribed AD molecules were venlafaxine (N= 14,064, 21.1%), paroxetine (N= 9,108, 13.6%), mirtazapine (N=7,478, 11.2%), escitalopram (N= 6,663, 10.0%) and fluoxetine (N=6,042, 9.0%).

**Table 3:**
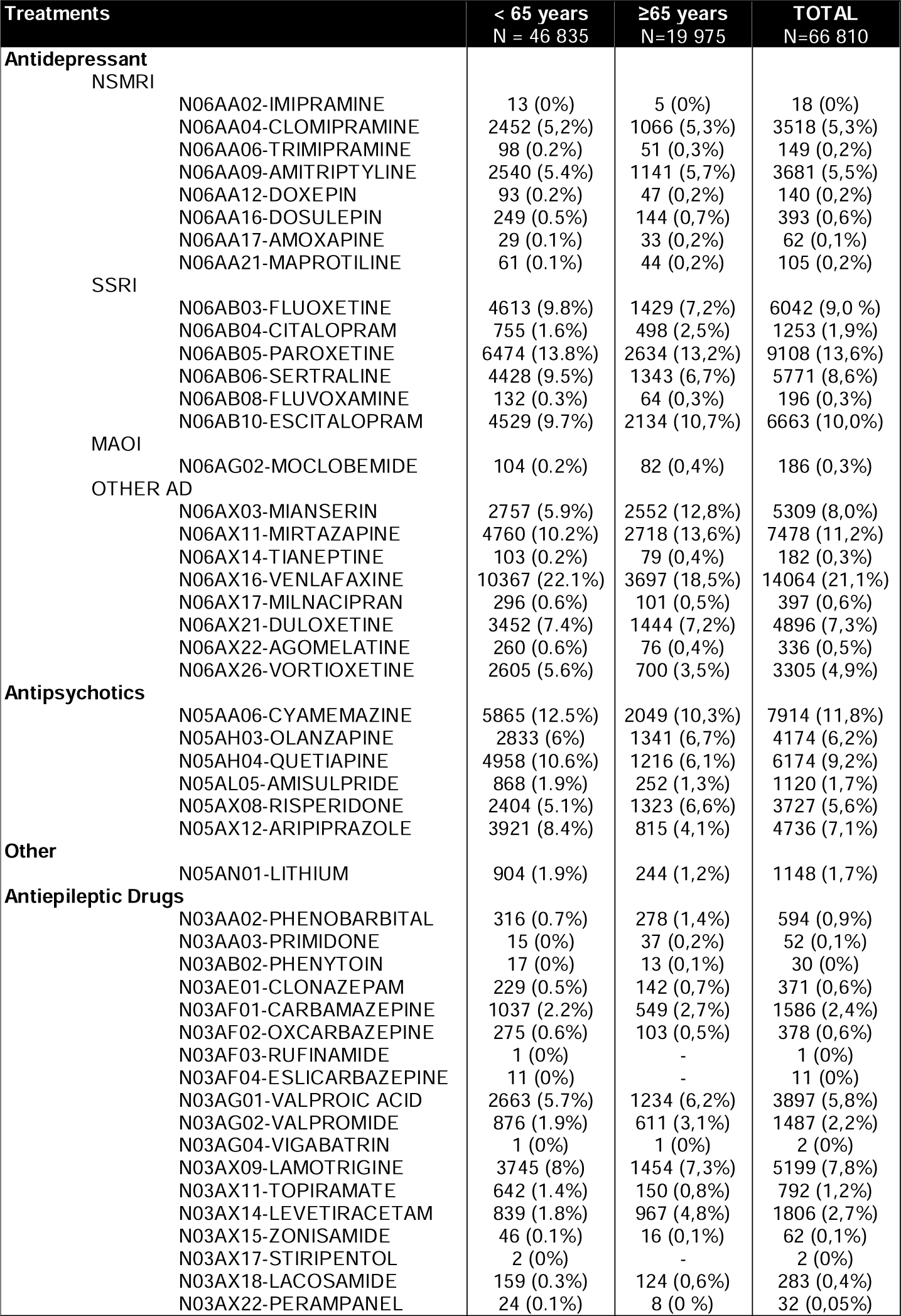
Molecules observed at last delivery (end of follow-up)

The most prescribed potentiators were quetiapine (n=6,174, 9.2%), followed by aripiprazole (n=4,736, 7.1%), olanzapine (n=4,174, 6.2%) and risperidone (n=3,727, 5.6%). Cyamemazine was also frequently used in TRD patients (n=7,914, 11,.8%).

Proportions across age groups were relatively similar, except for quetiapine, aripiprazole and venlafaxine that were less often prescribed in combination with an antidepressant among patients over 65 years than among younger ones (Quetiapine 10.6% vs 6.1%, Chi-square=229,75; p < 0.0001, Aripiprazole 8,4% vs 4,1%, Chi-square=322,5; p < 0.0001, Venlafaxine 22,1% vs 18,5%, Chi-square=154,75; p < 0.0001).

### Sensitivity analyses for prevalence estimation

The use of specific concomitant treatments (concomitant prescription) with an AD and/or a potentiator are presented in **Table 4**. Patients being prescribed any antidepressant that also has approval for anxiety disorders as well as any anxiolytic or cyamemazine were assumed to be possibly treated for an anxiety disorder. Patient being prescribed alcohol use disorder drugs or opioids concomitantly to their TRD strategy were assumed to be treated for alcohol use disorder and chronic pain respectively. Overall, 23.6% of TRD patients (n=15,759) were assumed to be treated for another psychiatric diagnosis, 14% (n=9,342) for an anxiety disorder, 8.6% (n=5,776) for neuropathic pain, and 2.5% (n=1,655) for alcohol use disorder. Excluding these patients, the annual prevalence of TRD was estimated at 25.8 per 10,000 persons in France.

**Table 4:** Patients probably treated for other primary diagnosis than MDD.

| **Strategy** | **< 65 years**<br>**N = 46 835** | **≥65 years**<br>**N = 19 975** | **TOTAL**<br>**N = 66 810** |
| --- | --- | --- | --- |
| **Patients probably treated for other primary diagnosis** | **11386 (24.3%)** | **4373 (21.9%)** | **15759 (23.6%)** |
| **Anxiety disorder** | **6790 (14.5%)** | **2552 (12.8%)** | **9342 (14%)** |
| AD with anxiety disorder indication (monotherapy) | 2725 (5.8%) | 1039 (5.2%) | 3764 (5.6%) |
| AD with anxiety disorder indication (mono) + anxiolytic | 3228 (6.9%) | 1269 (6.4%) | 4497 (6.7%) |
| ≥ 1 AD with anxiety disorder indication + cyamemazine | 249 (0.5%) | 71 (0.4%) | 320 (0.5%) |
| ≥ 1 AD with anxiety disorder indication + cyamemazine + anxiolytics | 588 (1.3%) | 173 (0.9%) | 761 (1.1%) |
| **Neuropathic pain** | **3848 (8.2%)** | **1928 (9.7%)** | **5776 (8.6%)** |
| AD + opioid | 812 (1.7%) | 424 (2.1%) | 1236 (1.9%) |
| ≥ 1 AD + POT + opioid | 3036 (6.5%) | 1504 (7.5%) | 4540 (6.8%) |
| **Alcohol use disorder** | **1496 (3.2%)** | **159 (0.8%)** | **1655 (2.5%)** |
| AD + alcohol use disorder drugs | 318 (0.7%) | 35 (0.2%) | 353 (0.5%) |
| ≥ 1 AD + POT + alcohol use disorder drugs | 1178 (2.5%) | 124 (0.6%) | 1302 (1.9%) |

Among the TRD population, most patients were prescribed anxiolytics/sedatives (55.8%, n=37,309), and a substantial proportion of them benefited from potentiated strategy (37.5%, n=25,036).

Proportions across age groups were relatively similar, with slightly higher prevalence of anxiety disorder and alcohol use disorder in patients above 65 years of age than in younger ones (anxiety disorder 14,5% vs 12,8%, Chi-square=2,13; p=0,14, alcohol use disorder 3,2% vs 0,8%, Chi-square=303,57; p < 0.0001, neuropathic pain 8,2% vs 9,7%, Chi-square=144,17; p < 0.0001).

## Discussion

To our knowledge, few population-based real-world studies have examined treatment patterns used by practitioners for adult patients with treatment-resistant depression. In this report, we aimed to estimate the prevalence of TRD and to provide evidence on treatment strategies from real-world data.

In a large sample of the French nationwide claims database, we found that the prevalence of TRD was 23.9%. This finding was slightly lower than the existing literature where the estimated prevalence of TRD ranged from 6.6% to 35% in patients with MDD (22,27–29), but based on different definitions of TRD. In a recent literature review (12), authors calculated the prevalence of TRD from the Sequenced Treatment Alternatives to Relieve Depression (STAR*D) trial (22) and estimated that approximatively 55% of patients with MDD did not respond to 2 treatment strategies within 12 weeks.

Estimates in our study corresponded to a sex- and age-standardized prevalence of 35.1 per 10,000 patients. When excluding patients potentially treated with AD for another reason than MDD, the annual prevalence was still high, at 25.8 per 10,000 persons. In France, according to last population-based studies in depression (3), the expected prevalence of TRD in France should be from 10 to 262 per 10 000 patients. In a similar study conducted from a sample of the French national claims database on a different time period (2012–2015), the prevalence was estimated to range between 25.8 and 37.6 per 10,000 patients (30).

Most patients were women (63.7%), especially in the patients over 65 years of age (72.1%), with median disease duration of 6 years, and median treatment duration for a TRD episode of 18 months. These findings were within the same range of studies conducted in patients with MDD identified from the French national healthcare claims (i.e. 56.7 years *versus* 54.6 years, and 64% *versus* 66%) (3).

The evaluation of treatment patterns for TRD revealed that combination of an AD and an antipsychotic was the most frequently used strategy (38,3%), followed by another AD monotherapy (31,9%). SSRIs were the AD the most prescribed in TRD whatever the strategy used, which is consistent with prior research (31–33). Of the potentiators, quetiapine, a second-generation antipsychotic, was the most prescribed. In Europe, quetiapine holds approval for being used as “add-on to ongoing antidepressant treatment for major depressive episodes in patients with MDD who have had sub-optimal response without remission to that treatment ADs” (34).

Benzodiazepines were found to be significantly prescribed in TRD patients (55.7%). This result was consistent with the existing literature (35). This recent cohort study found that benzodiazepines long-term use was over-prescribed in TRD patients (in almost a half of the patients) despite recommendations for withdrawal and the risk of worsening clinical and cognitive symptoms in this population (35). Our study strengthens previous findings that support progressive and planed withdrawal of benzodiazepines, particularly among TRD patients, and promote the most validate non-pharmacological alternatives when available (36,37).

This study has several strengths. The first relied on the high populational level of the database, which included all adult receiving an AD from 4 entire regions of France, accounting for 27% of the French individuals. Additionally, the SNDS database gathers all healthcare claims, allowing traceability of all reimbursed treatments.

Second, the TRD identification algorithm was developed to reflect the recent published recommendations of national experts (13). Although guidelines do not specify prioritization of strategies, our findings suggest some predominant stages. We found that 30% of TRD patients were treated with AD monotherapy, with lower duration of the disease (median duration of 4 years) compared to potentiated strategies (70%), more frequently used for longer disease durations (median duration of 7 years). This suggest that TRD patients tend to be treated with AD monotherapy at an early stage, while combining treatment with potentiator later on, especially with antipsychotics. Recently, a study in elderly TRD patients demonstrated that augmentation of existing ADs with aripiprazole over a short term period (10 weeks) improved well-being significantly than a switch to bupropion and was associated with higher remission (38), while continuing this type of treatment should be carefully considered since it can be at increased risk of mortality in patients with cognitive disorders. In this context, an expert-panel study conducted in the US evaluating adherence to guideline of primary care physicians pointed out the high need for improvement of treatment adjustment in nonresponsive patients, among other quality indicators (39).

Third, we provided estimation based on a more stringent definition to minimize overestimation of prevalence estimates. We intended to identify other potential primary diagnoses than MDD that a patient could be treated for with an AD-based strategy, such as anxiety disorders, chronic pain or alcohol use disorders, to exclude them from the prevalence calculation.

Our study has several limitations. First, the exact medical indication of the treatment is not clearly stated in the database, so that AD could have been used for another primary indication than MDD. Several conditions identified essentially through LTD and admissions were initially excluded from the overall population, such as bipolar disorder, chronic psychotic disorder, Parkinson’s disease and dementia, to confirm the underlying depressive condition. To reduce this risk, we performed sensitivity analyses, and we identified specific conditions that can be indications for ADs based on concomitant treatment deliveries, i.e. anxiety disorders, chronic pain and alcohol use disorder with comorbid depression. Unobserved other primary diagnoses can be confounding factors in a study carried out on a claims database which could lead to biased prevalence estimates. In our sensitivity analysis, primary indication for anxiety disorders was considered probable when an AD holding this indication was concomitantly prescribed with anxiolytics, or cyamemazine. When excluding these groups of patients, 23.6% of the TRD initial population, prevalence of TRD was estimated at 27.4 per 10,000 patients. In our study, we found that almost 7.8% of TRD patients were prescribed AD holding anxiety disorders indication with anxiolytics on top, which suggests that these patients may be potentially treated for anxious disorder as primary diagnosis. On top, 5.6% of patient were prescribed AD monotherapy holding anxiety disorder indication, and 1.6% with cyamemazine on top. In France, cyamemazine is especially used for its anxiolytic and sedative effect in treatment of depression.

Second, our definition of TRD was based on the sequence of treatment deliveries covering a period of 4 weeks, so that TRD patients hospitalized for a long period of time may have not been captured in our population. However, long-term hospitalized patients would have been captured with deliveries dispensed after a psychiatric hospitalization.

Another potential limitation was the minimum interval of 4 weeks required between two successive sequences of treatment strategy before considering that the treatment strategy may have failed. Although some studies may have used shorter interval, this assumption was based on the French guidelines which recommend waiting 4 to 6 weeks ideally before evaluating treatment efficacy even longer in older adults.

## Conclusion

This population-based study supports that TRD is prevalent in the French population and highlights the frequent use of second-generation antipsychotics as potentiator of AD medication (38,3%), followed by AD monotherapy (31,9%), in real-world settings. RCTs comparing these frequent therapeutic strategies are urgently needed to reduce the suffering and the major consequences caused by this disorder.

## Data Availability

Data are not publicly available

