## Supplementary for "Prevalence and National Patterns of Commonly Prescribed Antidepressants and other Psychotropic Medications to patients with treatment-resistant depression in France"

**Supplementary file 1 – List of code to identify disease and treatment of interest**

| **Pathology** | **ICD-10 codes for LTD and admission diagnoses** |
| --- | --- |
| Depression | F32, F33 |
| Psychotic disorders | F20-F29 |
| Bipolar affective disorder | F31 |
| Parkinson's disease | G20, F02.3 |
| Dementia | F00-F03, G30, F05.1 |
| **Treatments** | **ATC codes** |
| Antidepressants | Imipramine N06AA02  Clomipramine N06AA04  Trimipramine N06AA06  Amitriptyline N06AA09  Doxepin N06AA12  Dosulepin N06AA16  Amoxapine N06AA17  Maprotiline N06AA21  Fluoxetine N06AB03  Citalopram N06AB04  Paroxetine N06AB05  Sertraline N06AB06  Fluvoxamine N06AB08  Escitalopram N06AB10  Moclobemide N06AG02  Oxitriptan N06AX01  Mianserin N06AX03  Mirtazapine N06AX11  Tianeptine N06AX14  Venlafaxine N06AX16  Milnacipran N06AX17  Duloxetine N06AX21  Agomelatine N06AX22  Vortioxetine N06AX26 |
| Antipsychotics | Cyamemazine N05AH03  Olanzapine N05AH04  Quetiapine N05AL05  Amisulpride N05AN01  Risperidone N05AX12 |
| Other | Lithium N05AX08 |
| Antiepileptic drugs | Phenobarbital N03AA02  Primidone N03AA03  Phenytoin N03A02  Clonazepam N03AE01  Carbamazepine N03AF01  Oxcarbazepine N03AF02  Eslicarbazepine N03AF04  Valproic Acid N03AG01  Valpromide N03AG02  Lamotrigine N03AX09  Topiramate N03AX11  Levetiracetam N03AX14  Zonisamide N03AX15  Lacosamide N03AX18  Perampanel N03AX22 |
| **Anxiolytics and sedatives** |  |
| Benzodiazepines | Diazepam N05BA01  Chlordiazepoxide N05BA02  Oxazepam N05BA04  Potassium clorazepate N05BA05  Lorazepam N05BA06  Bromazepam N05BA08  Clobazam N05BA09  Prazepam N05BA11  Alprazolam N05BA12  Nordazepam N05BA16  Ethyl loflazepate N05BA18  Clotiazepam N05BA21 |
| H1-receptor antagonist | Hydroxyzine N05BB01  Hydroxyzine, combinations N05BB51 |
| Other anxyolitics | Tofisopam N05BA23  Mexazolam N05BA25  Captodiame N05BB02  Meprobamate N05BC01  Emylcamate N05BC03  Mebutamate N05BC04  Meprobamate, combinations N05BC51  Benzoctamine N05BD01  Buspirone N05BE01  Mephenoxalone N05BX01  Gedocarnil N05BX02  Etifoxine N05BX03  Fabomotizole N05BX04  Lavandulae aetheroleum N05BX05 |
| Alcohol use disorder drug | Disulfiram N07BB01  Calcium carbimide N07BB02  Acamprosate N07BB03  Naltrexone N07BB04  Nalmefene N07BB05 |
| Opioids | N02AA Natural opium alkaloids  N02AB Phenylpiperidine derivatives  N02AC Diphenylpropylamine derivatives  N02AD Benzomorphan derivatives  N02AE Oripavine derivatives  N02AF Morphinan derivatives  N02AG Opioids in combination with antispasmodics  N02AJ Opioids in combination with non-opioid analgesics  N02AX Other opioids |
| Thyroid hormones | Levothyroxine sodium H03AA01  Liothyronine sodium H03AA02  Combinations of levothyroxine and liothyronine H03AA03 |

**Supplementary File 2 – Illustrations of treatment strategies**

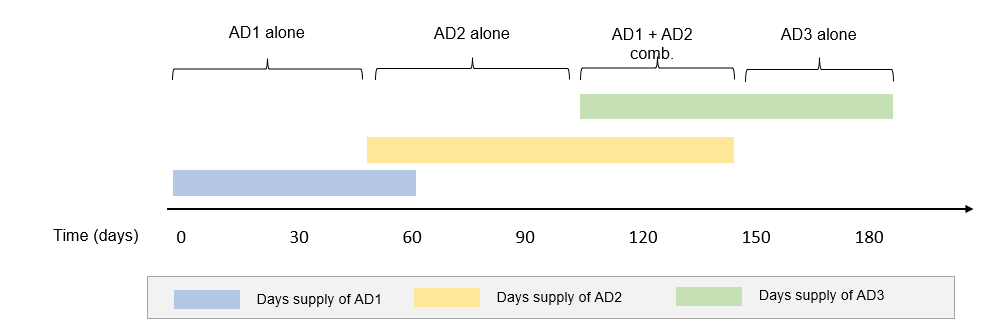

Figure 2 Illustration of initiation date for a treatment strategy

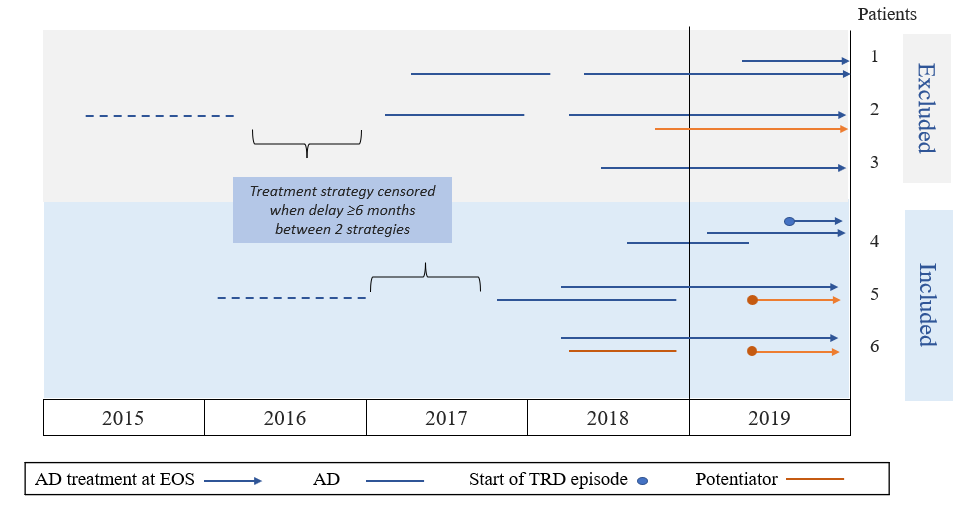

Figure 3 Illustration of patient inclusion and exclusion for each strategy

**Supplementary file 3 - Flow chart of included patients**

**Excluded patient with:**

Chronic psychotic disorder (N= 44,289, 11.1%)

Chronic Parkinson’s disease (N= 10,429, 2.6%)

Dementia (N= 37,185, 9.3%)

Bipolar disorder (N= 39,147, 9.8%)

**Adult patients with ≥1 delivery of AD in 2019 in the 4 regions**
N= 452,800

**TRD patients**

N= 66,810 (23.9%)

**Patients with ≥ 3 AD deliveries**
N= 279,479 (87.2%)

**Included patients for analyses**

N= 46,835 (70.1%) <65 years old

N= 19,975 (29.9%) ≥ 65 years old

**Excluded patient with:**

< 3 ADs deliveries in 2019: N= 40,886 (12.8%)

**Adult patients treated for depression**

N= 320,365 (70.7%)
